# Mapping the genetic landscape of the age at onset and severity of eating disorder symptoms

**DOI:** 10.1101/2025.09.25.25336612

**Authors:** Helena L. Davies, Abigail R. ter Kuile, Sang Hyuck Lee, Rujia Wang, Jessica Mundy, Zain-Ul-Abideen Ahmad, Jonathan Coleman, Saakshi Kakar, Emily Kelly, Chelsea Mika Malouf, Gursharan Kalsi, Moritz Herle, Gerome Breen, Christopher Hübel

## Abstract

**Background:** Eating disorder treatment is most successful when delivered at first onset, underscoring the importance of early intervention. UK patients transition from child to adult services at age 18, a process often poorly supported. Genetic factors may contribute to earlier onset, but molecular genetic research into symptom onset and severity remains limited.

**Aims:** We investigated (1) age at onset of behavioural symptoms and low weight, including sex differences, (2) the SNP-based heritability and cross-trait polygenic associations of symptoms, onset, and severity.

**Method:** Participants (n = 24,973) were from the NIHR BioResource, including the Genetic Links to Anxiety and Depression Study, Eating Disorders Genetics Initiative UK, and COVID-19 Psychiatry and Neurological Genetics Study. Symptoms were assessed with the ED100K questionnaire. We described sex-stratified onset of binge eating, low weight, self-induced vomiting, laxatives, diuretics, excessive exercise, and fasting. SNP-based heritability was estimated using GCTA-GREML. We tested associations with 26 polygenic scores (PGS), which were calculated using SBayesRC.

**Results:** Symptom median onset ranged from 16-20 years: vomiting (16), fasting and excessive exercise (17), binge eating (18), laxatives (19), diuretics (20), and low weight (20). Males were disproportionately likely to report adult onset. In females, behavioural symptoms and low weight showed substantial heritability (SNP-based *h*² = 0.31–0.78), but onset and cognitive symptoms appeared to be less heritable. PGS analyses identified distinct pathways: in females, higher childhood obesity PGS was linked to earlier binge eating onset (∼7 months), and higher educational attainment PGS to earlier low weight onset (∼6 months).

**Conclusions:** Eating disorder symptoms begin in adulthood as frequently as adolescence, challenging stereotypes and underscoring investment needed in adult services, particularly for men. While symptoms were strongly heritable, onset and cognitive symptoms showed limited genetic contribution. Symptom-level PGS analyses highlight distinct biological and developmental pathways. Larger sample sizes and more accurate phenotyping are needed.

## Introduction

Eating disorders affect at least ∼1.4 million people in the UK (1). The most-studied are: anorexia nervosa, which presents as severe weight loss (BMI <18.5) due to restriction or excessive exercise with a perception of fatness despite low weight (2); bulimia nervosa, which involves binge eating followed by compensatory behaviours such as self-induced vomiting; and binge-eating disorder, which presents as binge eating without compensation (2). Treatment outcomes for eating disorders are poor (3–6), but interventions are most successful when delivered at first onset (7). Thus, early intervention is important (8).

### Age at onset of eating disorders and treatment in the UK

Early identification should target the transition from adolescence into emerging adulthood as this is when eating disorders typically begin. A meta-analysis (k=192) summarised retrospectively reported median age at symptom onset (7): anorexia nervosa was 16 years, bulimia nervosa was 18 years, and binge-eating disorder was 20 years (9–11). No UK samples were included, which we address with our study. In the UK, authorities require treatment to be delivered in age-appropriate settings (12) and patients turning 18 years old move to adult services. This transition lacks structure and is complicated by changes in treatment, social environment, and family structure (12). A 2019 National Health Service (NHS) England Guidance called for a seamless transition pathway (13). Moreover, eating disorder symptoms differ by onset; compared to an adolescent onset (13–20 years), children (<13 years) report less binge eating, purging, or excessive exercise for weight loss (14,15) and lose weight at a faster pace (14). Adolescent onset of anorexia nervosa shows lower death rates and better long-term outcomes than later onset (16), whilst onset in childhood shows worse outcomes (17,18).

### Genetics of eating disorders and their age at onset

Genetic risk may influence age at onset, as eating disorder symptoms are heritable (**Table 1**) and early onset may reflect greater genetic susceptibility (19–24). The Brainstorm Consortium reported a negative correlation between average age of onset and heritability across 22 brain disorders, including anorexia nervosa (25). For instance, Tourette’s syndrome showed 20% heritability with an average onset of 7 years whilst major depressive disorder showed 13% heritability with an average onset of 32 years (25). Adult-onset ADHD is associated with a lower ADHD polygenic score than childhood-onset, suggesting distinct subtypes, with the former being more environmentally influenced, less consistent, or milder (26). Sex-specific genetics may also play a role in onset: early-onset schizophrenia in females may reflect greater genetic susceptibility, whilst males may accumulate more environmental risks (19). Such findings emphasise how genetic risk interacts with early environment and sex-at-birth.

**Table 1.** Twin-based heritability of eating disorder symptoms and age at diagnosis of anorexia nervosa.

|  | **Twin-based heritability (Reference)** |
| --- | --- |
| **Anorexia nervosa symptoms** |  |
| Intentional weight loss (males) | 38% (27) |
| Intentional weight loss (females) | 66% (27) |
| Low weight (BMI < 18.5; females) | 31% (28) |
| ‘Dieting’ factor | 42% (29) |
| Afraid of weight gain during low weight | 27% (28) |
| Feeling fat at low weight | 23% (28) |
| Undue influence of weight on self-esteem at low weight | 29% (28) |
| Drive for thinness (females) | 51% (30) |
|  | 44% (29) |
| Drive for thinness (males) | 0% (30) |
| **Bulimia nervosa symptoms** |  |
| Self-induced vomiting (females) | 72% (31) |
|  | 53% (32) |
| Laxatives (females) | 43% (32) |
| Diuretics (females) | 44% (32) |
| Diet pills (females) | 51% (32) |
| **Binge-eating disorder symptoms** |  |
| Binge eating in the absence of compensatory behaviours (males & females) | 41% (33) |
| **Transdiagnostic symptoms: AN and BN** |  |
| Fasting (females) | 42% (32) |
| Excessive exercise (females) | 35% (32) |
| Body dissatisfaction (females) | 59% (30) |
|  | 52% (29) |
| Body dissatisfaction (males) | 0% (29,30) |
| Weight and shape concern (females) | 25% (34) |
| Importance of weight/shape to self-evaluation (females) | 24% (32) |
| **Transdiagnostic symptoms: BN and BED** |  |
| Binge eating (females) | 34% (35) |
|  | 46% (31) |
|  | 34% (32) |
|  | 49% (36) |
|  | 70% (37) |
| Binge eating (males) | 74% (37)* |
| Binge eating (males & females) | 51% (38) |
| **Age at diagnosis (GCTA GREML)** |  |
|  | **SNP-based**<br>**heritability**<br>**(Reference)** |
| Anorexia nervosa (continuous) | 1% (39) |
| Early-onset anorexia nervosa (i.e., binary diagnosis under the age of 13 and assuming lifetime prevalence of 0.1%) | 21% (39) |
| Typical-onset anorexia nervosa (i.e., diagnosis over the age of 13 and assuming lifetime prevalence of 0.9%) | 15% (39) |

\* The authors noted that they were only able to estimate univariate heritabilities due to small sample sizes.
*Note.* We have only linked symptoms to the following three eating disorders: anorexia nervosa, bulimia nervosa, and binge-eating disorder. However we would like to acknowledge that these symptoms appear in other eating disorders such as purging disorder.

Genome-wide complex trait analysis via genomic-relatedness-based restricted maximum-likelihood (GCTA-GREML) of the age of diagnosis of anorexia nervosa showed a SNP-based heritability of 1%, 21% for early-onset (diagnosis <13 years), and 15% for typical-onset (>13 years, **Table 1**), suggesting that earlier onset may be driven by genetics (39). Early onset showed a negative genetic correlation with age at menarche, and Mendelian randomisation suggested a causal role of younger age at menarche on early-onset anorexia nervosa (39). These findings indicate distinct sub-phenotypes influenced by puberty timing. An age at menarche polygenic score also correlated negatively with binge-eating disorder (40) and positively with adolescent persistent thinness (41), suggesting broader relevance for eating disorders and weight phenotypes beyond anorexia nervosa.

### Aims

As of 2025, the genetics of eating disorder symptoms and their clinical course, including onset and severity, have been minimally investigated. Therefore, we performed a two-fold investigation: 1) phenotypic analysis exploring the age at onset of behavioural symptoms - including binge eating, self-induced vomiting, diuretic and laxative use, fasting, and excessive exercise - and low weight in a large UK sample; 2) a genetic analysis to assess the SNP-based heritability and cross-trait polygenic risk score association of a) behavioural eating disorder symptoms and low weight and b) their age at onset, and c) cognitive symptoms and d) their severity.

## Methods

### Sample

Participants were from the National Institute for Health and Care Research (NIHR) BioResource (*n* = 70,442) who participated in the Genetic Links to Anxiety and Depression (GLAD) Study (42) (*n* = 46,802), the Eating Disorders Genetics Initiative (43) (EDGI UK; *n* = 6,393), or other physical health and general population cohorts (*n* = 17,247) whose participants took part in the COVID-19 Psychiatry and Neurological Genetics (COPING) Study (**Supplementary Table 1**). The COPING Study provided complementary data for GLAD and EDGI participants to their sign-up survey (44).

We excluded 45,469 participants missing sex (*n* = 1,826), age (*n* = 1,474), or eating disorder symptoms (*n* = 41,321). The ED100K was optional in GLAD, which explains the high missingness. We excluded 1,322 participants with IBS/IBD to match the general population’s illness prevalence. 24,973 individuals reportedat least one eating disorder symptom, whilst 16,846 had age at onset data. We excluded 337 participants with genetically-inferred non-European ancestry via principal components analysis. We removed genetic variants with minor allele frequency <1%, missing in >5% of participants, and Hardy-Weinberg equilibrium deviation (*p* < 10^-10^). We excluded participants with >5% missing variants, undetermined genetic sex, or identity-by-descent outliers indicating contamination across genetic samples (*n* = 409), leaving 15,559 individuals with both phenotypes and genetic data. For the PGS analysis, we removed UK Biobank participants (*n* = 1,132) to avoid sample overlap with discovery genome-wide association studies (GWASs) conducted within the UK Biobank.

### Ethical approval

The authors assert that all procedures contributing to this work comply with the ethical standards of the relevant national and institutional committees on human experimentation and with the Helsinki Declaration of 1975, as revised in 2013. The London - Fulham Research Ethics Committee approved the GLAD Study, 21.08.2018 (REC reference: 18/LO/1218) and EDGI UK on 29.07.2019 (REC reference: 19/LO/1254). The NIHR BioResource is approved as a Research Tissue Bank by the East of England - Cambridge Central Committee (REC reference: 17/EE/0025). The COPING study is approved by the South West - Central Bristol Research Ethics Committee on 27.04.2020 (REC reference: 20/SW/0078). All participants provided informed consent.

### Measures

#### Behavioural eating disorder symptoms and low weight

Participants reported binge eating, low weight, self-induced vomiting, laxative use, diuretic use, fasting, and excessive exercise via the ED100K (45) (**Supplementary Table 2**). This questionnaire was included in the sign-up survey in EDGI UK and COPING and was optional in GLAD. We defined ‘cases’ as participants who reported a lifetime symptom and ‘controls’ as participants who endorsed not having experienced that symptom and who were not recruited on the basis of having a psychiatric disorder.

Participants endorsing a symptom could report the age at onset (**Supplementary Table 2**): for example, “*Roughly how old were you when you began having regular episodes of binge eating?*”. For binge eating, participants must endorse loss of control (*n*_excluded_ = 13). We removed implausible responses younger than 3 and older than 118 years (*n*_excluded_ = 257) and ages that exceeded the participant’s current age (*n*_excluded_ = 58).

#### Cognitive symptoms

Participants reported the severity of five cognitive symptoms on Likert scales: 1) self-worth being dependent on body shape or weight when at low weight and 2) when binge eating, 3) fear of weight gain at low weight, 4) feeling fat at low weight, and 5) feeling that body parts are larger than they are at low weight **(Supplementary Table 2).** We generated binary (symptom present or absent) and continuous (severity scores) variables for all symptoms except ‘feeling that body parts are larger’, which we retained as a 3-point scale (“Not at all”, “Somewhat”, “Very much”). For the remaining continuous variables, we excluded participants who endorsed absence of the symptom, creating severity scales ranging from 4 to 6 points (depending on the symptom) that reflect the range in those who with the symptom.

#### Analysis

We used R version 4.1.2 (46).

##### Retest-reliability of retrospective self-reported age at onset

A small sub-sample of participants completed the ED100K twice: first, optionally in the GLAD Study and second, mandatorily in the COPING study (*n* = 239 to 1,805 participants, depending on symptom). We assessed participants’ reliability of their retrospective reports by calculating the difference between both answers. For our analyses, we used participants’ most recent responses.

##### Phenotypic analysis

**Age at onset.** We described combined and sex-stratified age at onset. We calculated percentages in onset categories by sex: (1) Childhood: ≤12 years, (2) Adolescence: 13–17 years, (3) Emerging adulthood (47): 18–25 years, and (4) Adulthood: ≥26 years.

### Genetic analysis

#### SNP-based heritability

Using GCTA-GREML (48), we estimated SNP-based heritabilities in females only (insufficient male cases, *n*_max_ = 665). Population prevalences for liability-scale estimation of behavioural symptoms and low weight were estimated from the part of the sample not ascertained for a psychiatric disorder. We used our sample’s prevalence estimates for the cognitive symptoms, as they are likely to best reflect the population prevalence given that they exist only in cases (e.g., feeling fat *at low weight*). We estimated SNP-based *h^2^* for 1) quantitative and 2) binary age at onset, split into adult (≥18 years) and child/adolescent onset (<18 years). Quantitative traits were right-skewed and thus square-root transformed to approximate normality (**Supplementary Figure 1**). We assessed cognitive symptom severity using linear models, assuming an underlying latent normal distribution.

Analyses were adjusted for genotyping batches and ten genetic principal components. As age is correlated with differences in reporting onset between the GLAD and COPING studies (r = 0.23–0.29), we controlled for age in an additional analysis of onset.

#### Cross-trait polygenic associations

We created psychiatric, anthropometric, social, behavioural, somatic, and personality polygenic risk scores (PGS) (for discovery GWASs, see **Supplementary Table 3**) using SBayesRC(49). Using linear and logistic (binary and ordinal) regressions, we used these PGSs as exposure variables and explored their associations with the following outcomes: 1) behavioural symptoms and low weight, 2) their age at onset, 3) cognitive symptoms and 4) their severity in both females and males (where sample size allowed). PGSs were standardised. We used the same adjustment variables as above. We corrected for multiple testing using FDR with a 0.05 threshold.

## Results

### Sample descriptives

Participants had an average age of 34-39 years, largely female (89–96%), white (94–95%), and over 10% had a Master’s degree or above (**Supplementary Table 4**). For descriptive statistics of participants with both genetic data and behavioural symptoms and low weight, onset, cognitive symptoms, or cognitive severity data, see **Supplementary Tables 5–8 (females)** and **9-12 (males)**.

### Retest-reliability of retrospective self-reported age at onset

Binge eating showed the largest discrepancies in reported age at onset between first and second reporting in the GLAD and COPING studies, with 12% of the differences exceeding 10 years compared with 3.1% to 8.5% for the other eating disorder symptoms (**Supplementary Table 13; Supplementary Figure 2**). The mean and median differences ranged from 2 to 4.4 years and 1 to 2 years, respectively.

### Phenotypic analysis

**Age at onset.** Self-induced vomiting had the youngest median onset at 16 years in females (**Table 2; Supplementary Figure 3**). Fasting and laxative use in males had the oldest median onset at 30 years. Males consistently showed an older median age at onset than females across all symptoms; for example, the median age at onset of binge eating and low weight was 18 and 19 years in females but 22 and 24 years in males, respectively.

**Table 2.**
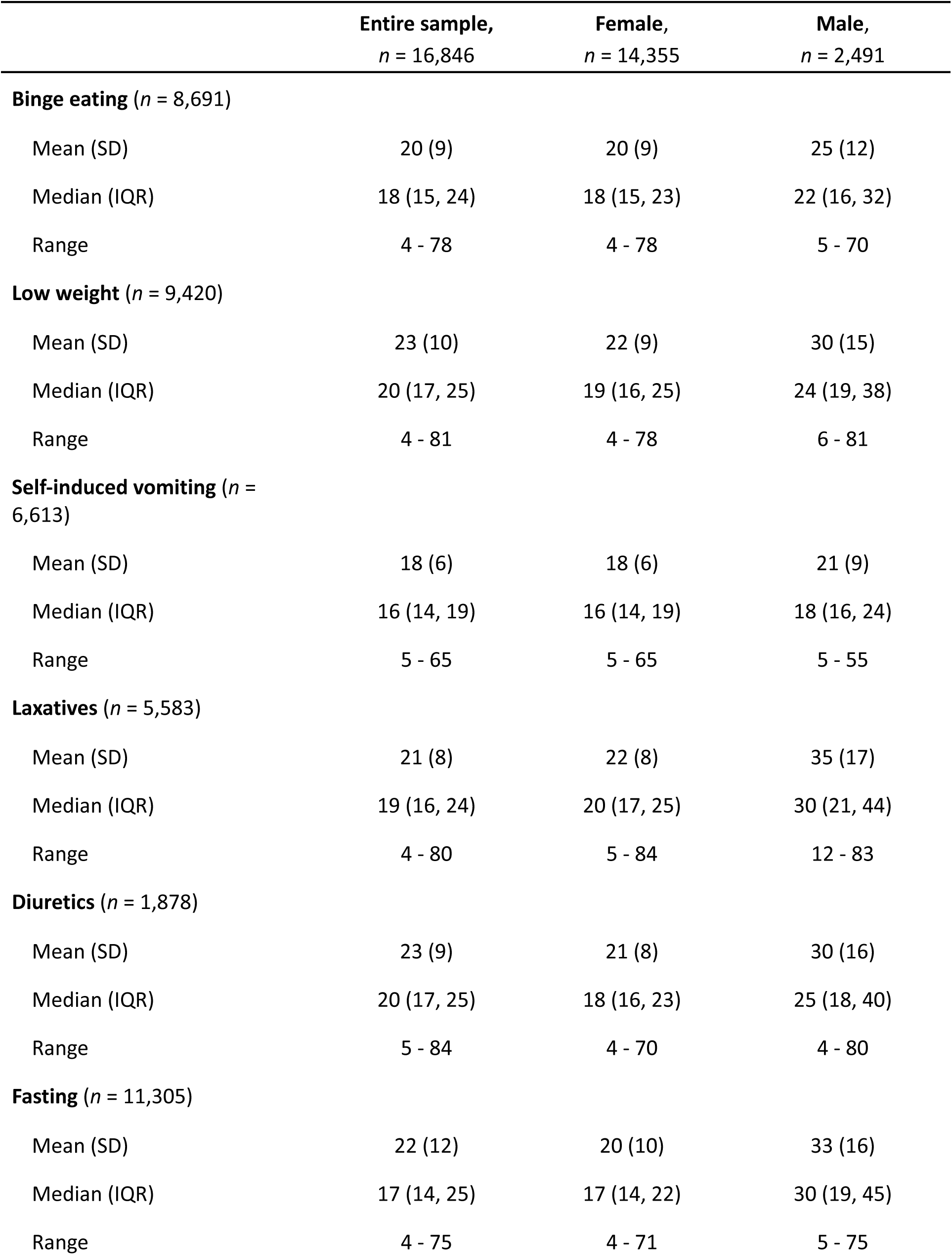
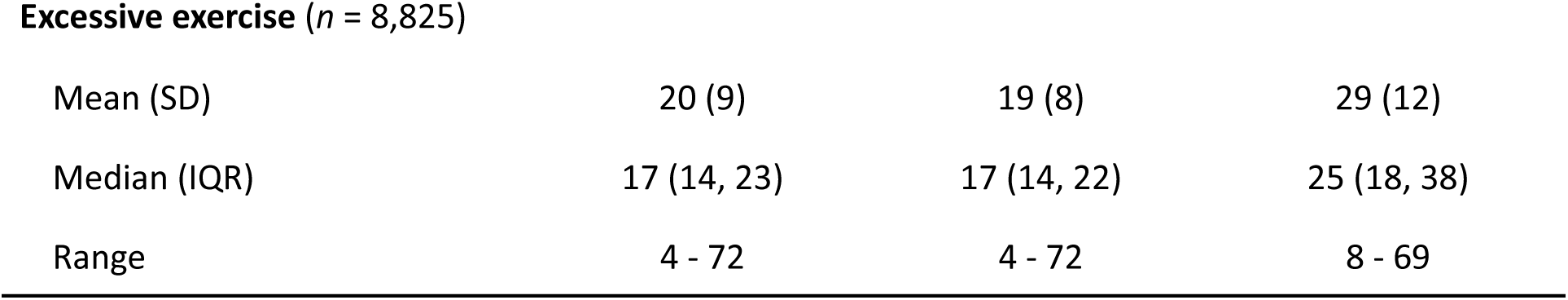
Mean, median, and range of age at onset of each phenotype in the entire sample with non-missing onset data, split by sex.

|  | Entire sample,<br>*n* = 16,846 | Female,<br>*n* = 14,355 | Male,<br>*n* = 2,491 |
| --- | --- | --- | --- |
| **Binge eating** (*n* = 8,691) |  |  |  |
| Mean (SD) | 20 (9) | 20 (9) | 25 (12) |
| Median (IQR) | 18 (15, 24) | 18 (15, 23) | 22 (16, 32) |
| Range | 4 - 78 | 4 - 78 | 5 - 70 |
| **Low weight** (*n* = 9,420) |  |  |  |
| Mean (SD) | 23 (10) | 22 (9) | 30 (15) |
| Median (IQR) | 20 (17, 25) | 19 (16, 25) | 24 (19, 38) |
| Range | 4 - 81 | 4 - 78 | 6 - 81 |
| **Self-induced vomiting** (*n* = 6,613) |  |  |  |
| Mean (SD) | 18 (6) | 18 (6) | 21 (9) |
| Median (IQR) | 16 (14, 19) | 16 (14, 19) | 18 (16, 24) |
| Range | 5 - 65 | 5 - 65 | 5 - 55 |
| **Laxatives** (*n* = 5,583) |  |  |  |
| Mean (SD) | 21 (8) | 22 (8) | 35 (17) |
| Median (IQR) | 19 (16, 24) | 20 (17, 25) | 30 (21, 44) |
| Range | 4 - 80 | 5 - 84 | 12 - 83 |
| **Diuretics** (*n* = 1,878) |  |  |  |
| Mean (SD) | 23 (9) | 21 (8) | 30 (16) |
| Median (IQR) | 20 (17, 25) | 18 (16, 23) | 25 (18, 40) |
| Range | 5 - 84 | 4 - 70 | 4 - 80 |
| **Fasting** (*n* = 11,305) |  |  |  |
| Mean (SD) | 22 (12) | 20 (10) | 33 (16) |
| Median (IQR) | 17 (14, 25) | 17 (14, 22) | 30 (19, 45) |
| Range | 4 - 75 | 4 - 71 | 5 - 75 |
| **Excessive exercise (*n* = 8,825)** |  |  |  |
| Mean (SD) | 20 (9) | 19 (8) | 29 (12) |
| Median (IQR) | 17 (14, 23) | 17 (14, 22) | 25 (18, 38) |
| Range | 4 - 72 | 4 - 72 | 8 - 69 |

Females more frequently reported adolescent onset (35–53%) than males (12–34%), whilst males more frequently reported adult onset (25–71%) than females (9–32%) (**Figure 1; Supplementary Table 14**). Further, more females than males reported onset in childhood of excessive exercise (16% vs. 2%) and fasting (16% vs. 4%).

**Figure 1.**
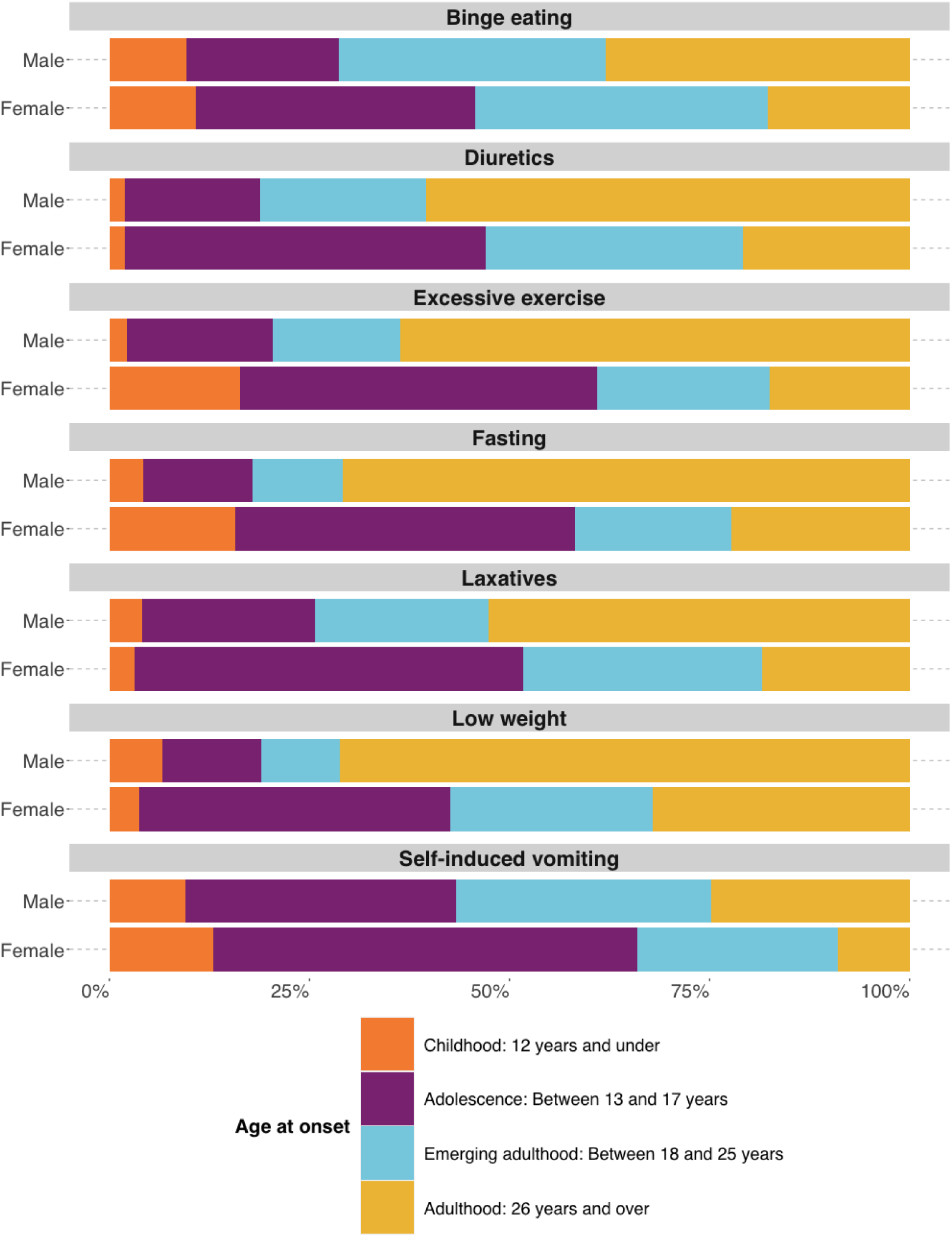
Sex-stratified age at onset split into four categories: 1) Childhood, 2) Adolescence, 3) Emerging adulthood, 4) Adulthood.

### Genetic analysis

#### SNP-based heritability

We detected significant SNP-based heritability for all behavioural symptoms and low weight (**Table 3**). Diuretic use had the highest heritability estimate (0.78 [SE = 0.10]) whilst low weight was the least heritable with (0.31 [SE = 0.08]).

**Table 3.**
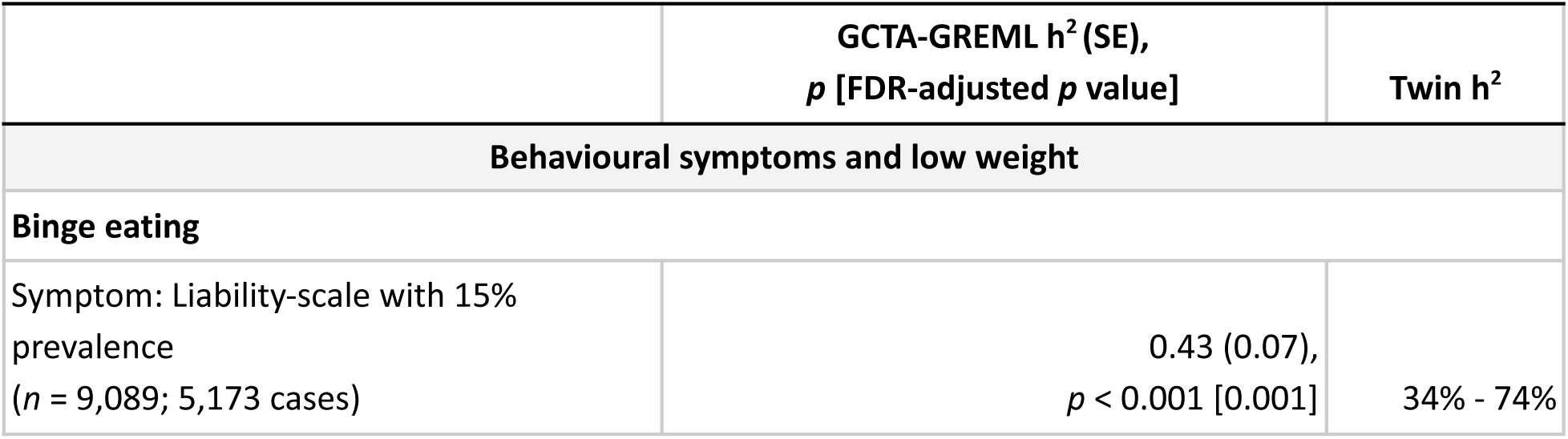
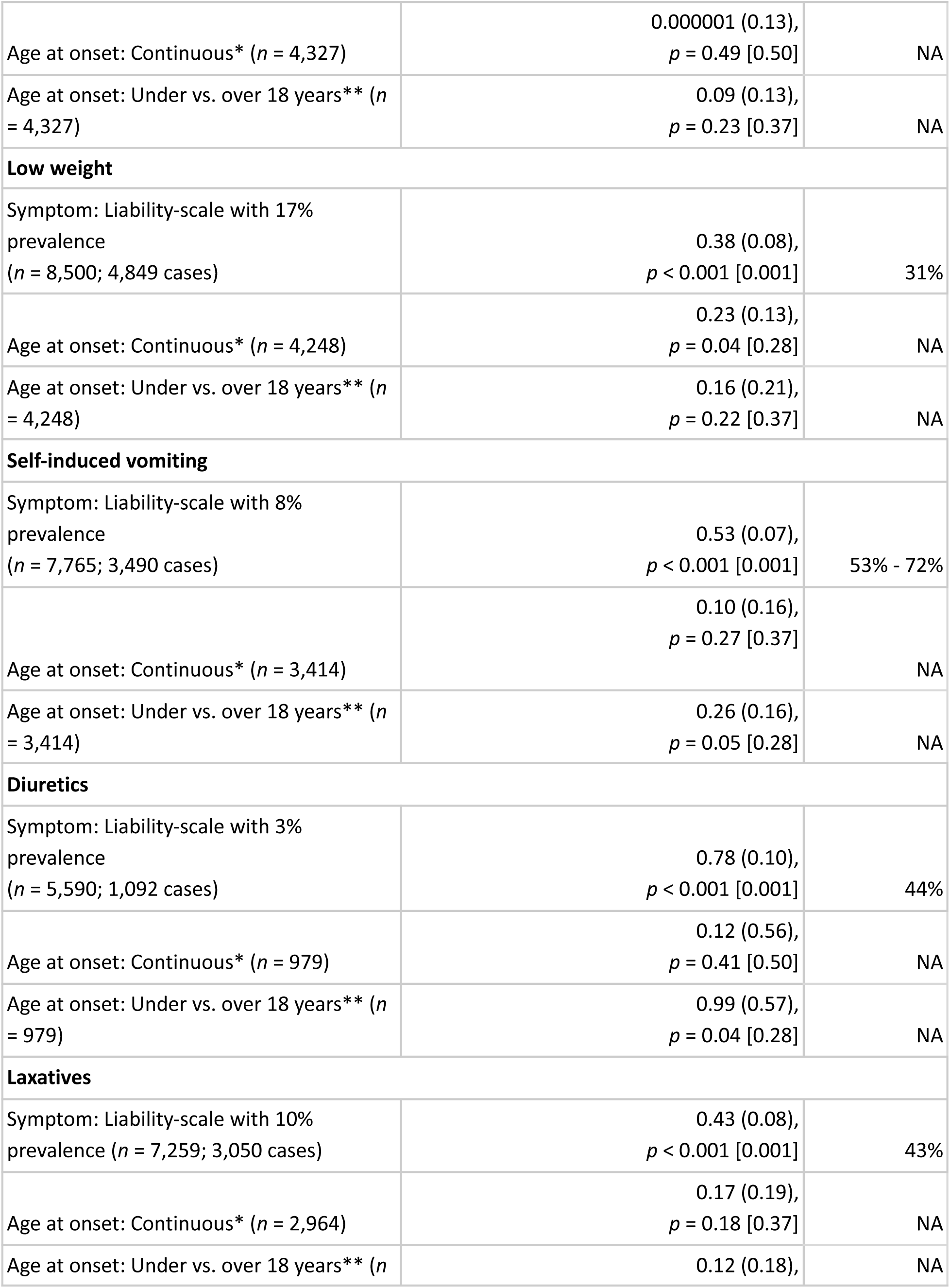
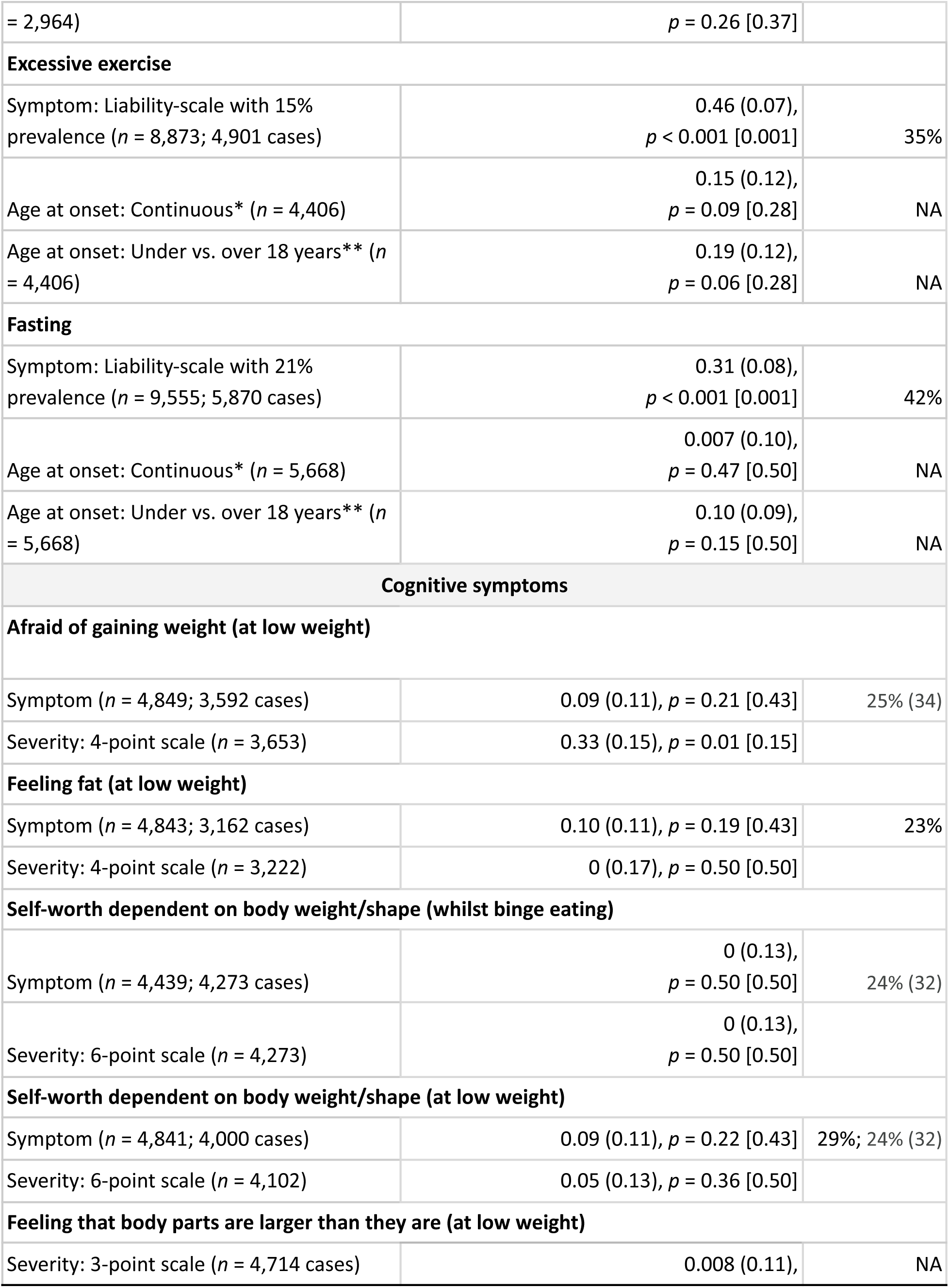
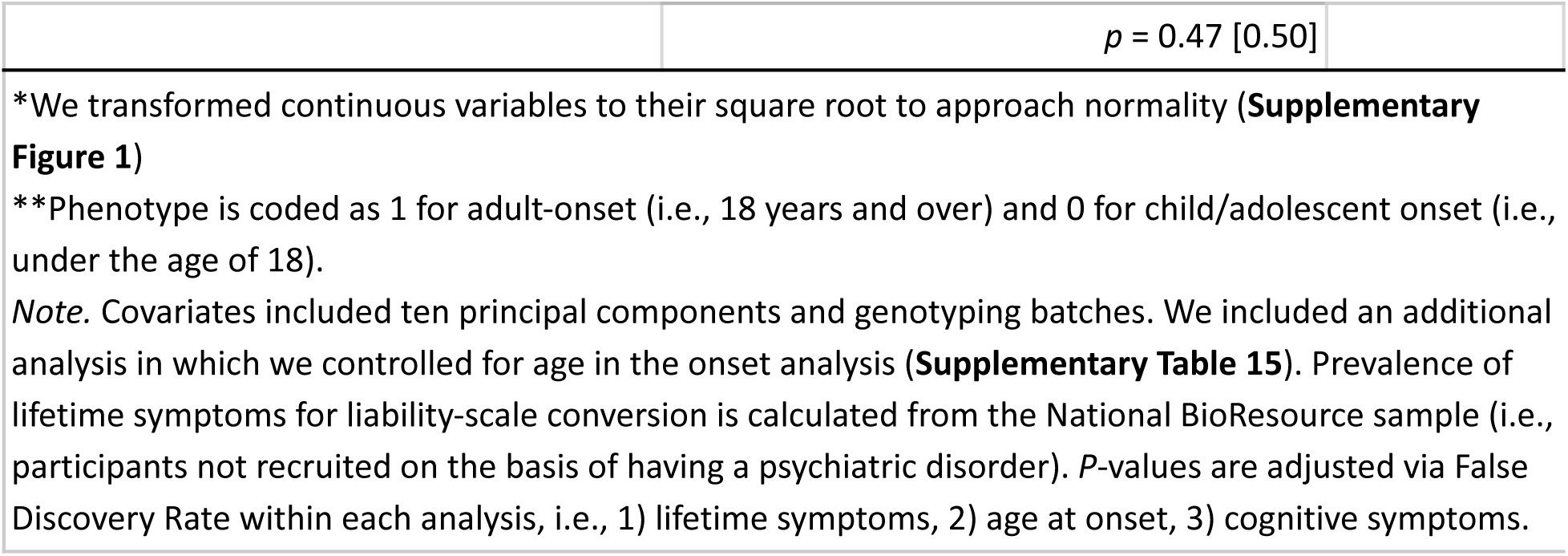
GCTA-GREML heritability estimates in females of lifetime presence and age at onset of binge eating, low weight, self-induced vomiting, diuretics, laxatives, excessive exercise, fasting, and cognitive symptoms and their severity.

| | GCTA-GREML $h^2$ (SE),<br>$p$ [FDR-adjusted $p$ value] | Twin $h^2$ |
| --- | --- | --- |
| **Behavioural symptoms and low weight** |  |  |
| **Binge eating** |  |  |
| Symptom: Liability-scale with 15% prevalence<br>($n = 9,089$; 5,173 cases) | 0.43 (0.07),<br>$p < 0.001$ [0.001] | 34% - 74% |
| Age at onset: Continuous* (*n* = 4,327) | 0.000001 (0.13),<br>*p* = 0.49 [0.50] | NA |
| Age at onset: Under vs. over 18 years** (*n* = 4,327) | 0.09 (0.13),<br>*p* = 0.23 [0.37] | NA |
| **Low weight** |  |  |
| Symptom: Liability-scale with 17% prevalence<br>(*n* = 8,500; 4,849 cases) | 0.38 (0.08),<br>*p* < 0.001 [0.001] | 31% |
| Age at onset: Continuous* (*n* = 4,248) | 0.23 (0.13),<br>*p* = 0.04 [0.28] | NA |
| Age at onset: Under vs. over 18 years** (*n* = 4,248) | 0.16 (0.21),<br>*p* = 0.22 [0.37] | NA |
| **Self-induced vomiting** |  |  |
| Symptom: Liability-scale with 8% prevalence<br>(*n* = 7,765; 3,490 cases) | 0.53 (0.07),<br>*p* < 0.001 [0.001] | 53% - 72% |
| Age at onset: Continuous* (*n* = 3,414) | 0.10 (0.16),<br>*p* = 0.27 [0.37] | NA |
| Age at onset: Under vs. over 18 years** (*n* = 3,414) | 0.26 (0.16),<br>*p* = 0.05 [0.28] | NA |
| **Diuretics** |  |  |
| Symptom: Liability-scale with 3% prevalence<br>(*n* = 5,590; 1,092 cases) | 0.78 (0.10),<br>*p* < 0.001 [0.001] | 44% |
| Age at onset: Continuous* (*n* = 979) | 0.12 (0.56),<br>*p* = 0.41 [0.50] | NA |
| Age at onset: Under vs. over 18 years** (*n* = 979) | 0.99 (0.57),<br>*p* = 0.04 [0.28] | NA |
| **Laxatives** |  |  |
| Symptom: Liability-scale with 10% prevalence (*n* = 7,259; 3,050 cases) | 0.43 (0.08),<br>*p* < 0.001 [0.001] | 43% |
| Age at onset: Continuous* (*n* = 2,964) | 0.17 (0.19),<br>*p* = 0.18 [0.37] | NA |
| Age at onset: Under vs. over 18 years** (*n* | 0.12 (0.18), | NA |
| = 2,964) | $p = 0.26 [0.37]$ | |
| **Excessive exercise** |  |  |
| Symptom: Liability-scale with 15% prevalence ($n = 8,873$; 4,901 cases) | 0.46 (0.07),<br>$p < 0.001 [0.001]$ | 35% |
| Age at onset: Continuous* ($n = 4,406$) | 0.15 (0.12),<br>$p = 0.09 [0.28]$ | NA |
| Age at onset: Under vs. over 18 years** ($n = 4,406$) | 0.19 (0.12),<br>$p = 0.06 [0.28]$ | NA |
| **Fasting** |  |  |
| Symptom: Liability-scale with 21% prevalence ($n = 9,555$; 5,870 cases) | 0.31 (0.08),<br>$p < 0.001 [0.001]$ | 42% |
| Age at onset: Continuous* ($n = 5,668$) | 0.007 (0.10),<br>$p = 0.47 [0.50]$ | NA |
| Age at onset: Under vs. over 18 years** ($n = 5,668$) | 0.10 (0.09),<br>$p = 0.15 [0.50]$ | NA |
| **Cognitive symptoms** |  |  |
| **Afraid of gaining weight (at low weight)** |  |  |
| Symptom ($n = 4,849$; 3,592 cases) | 0.09 (0.11), $p = 0.21 [0.43]$ | 25% (34) |
| Severity: 4-point scale ($n = 3,653$) | 0.33 (0.15), $p = 0.01 [0.15]$ | |
| **Feeling fat (at low weight)** |  |  |
| Symptom ($n = 4,843$; 3,162 cases) | 0.10 (0.11), $p = 0.19 [0.43]$ | 23% |
| Severity: 4-point scale ($n = 3,222$) | 0 (0.17), $p = 0.50 [0.50]$ | |
| **Self-worth dependent on body weight/shape (whilst binge eating)** |  |  |
| Symptom ($n = 4,439$; 4,273 cases) | 0 (0.13),<br>$p = 0.50 [0.50]$ | 24% (32) |
| Severity: 6-point scale ($n = 4,273$) | 0 (0.13),<br>$p = 0.50 [0.50]$ | |
| **Self-worth dependent on body weight/shape (at low weight)** |  |  |
| Symptom ($n = 4,841$; 4,000 cases) | 0.09 (0.11), $p = 0.22 [0.43]$ | 29%; 24% (32) |
| Severity: 6-point scale ($n = 4,102$) | 0.05 (0.13), $p = 0.36 [0.50]$ | |
| **Feeling that body parts are larger than they are (at low weight)** |  |  |
| Severity: 3-point scale ($n = 4,714$ cases) | 0.008 (0.11), | NA |

|  |  |
| --- | --- |
| | $p = 0.47 [0.50]$ |
| *We transformed continuous variables to their square root to approach normality (**Supplementary Figure 1**)<br>**Phenotype is coded as 1 for adult-onset (i.e., 18 years and over) and 0 for child/adolescent onset (i.e., under the age of 18).<br>*Note.* Covariates included ten principal components and genotyping batches. We included an additional analysis in which we controlled for age in the onset analysis (**Supplementary Table 15**). Prevalence of lifetime symptoms for liability-scale conversion is calculated from the National BioResource sample (i.e., participants not recruited on the basis of having a psychiatric disorder). *P*-values are adjusted via False Discovery Rate within each analysis, i.e., 1) lifetime symptoms, 2) age at onset, 3) cognitive symptoms. |  |

Neither the continuous nor the binary age at onset phenotypes showed a significant heritable component after correcting for multiple testing (**Table 3**) or adjusting for age (**Supplementary Table 15**). GCTA-GREML power calculations (50) indicated that our smallest sample (n_diuretic use_ = 979) translated to 9% power to detect a heritability of 0.20. Our largest sample (n_fasting_ = 5,668) had 95% power to detect a heritability >0.2, suggesting that the maximum heritability of this symptom’s continuous age at onset phenotype is <0.20. With this phenotype, we had 15% power to detect a heritability of 0.05, as was found for depression by Harder et al (2022).

We did not identify heritable components for cognitive symptoms or their severity at the current sample size (**Table 3**). Power calculations indicated that our largest sample sizes (*n*_symptom_ = 4,849 and *n*_severity_ = 4,714) were 100% powered to detect a heritability of 0.50 and 0.30, respectively.

#### Cross-trait polygenic associations

We observed multiple consistent significant associations across the behavioural symptoms and low weight in females (**Figure 2; Supplementary Table 16**) and males (**Supplementary Figure 4; Supplementary Table 17**). For example, the ADHD PGS was associated with greater odds of all symptoms in females (ORs = 1.12–1.36, *q*s < 0.001) and with binge eating, fasting, and excessive exercise in males (ORs =1.15–1.35, *q*s < 0.03). Other PGSs showed diverging associations. For example, in females the childhood obesity PGS was associated with lower odds of low weight (OR=0.85, 95% CI 0.8, 0.91, *q* < 0.001) but greater odds of both binge eating (OR=1.14, 95% CI 1.05, 1.24, *q* = 0.003) and excessive exercise (OR=1.10, 95% CI 1.02, 1.19, *q* = 0.022), and showed no significant association with any purging behaviour or fasting. For example, for each standard deviation (SD) increase in the childhood obesity PGS, the odds of reporting low weight were lower by 0.15.

**Figure 2.**
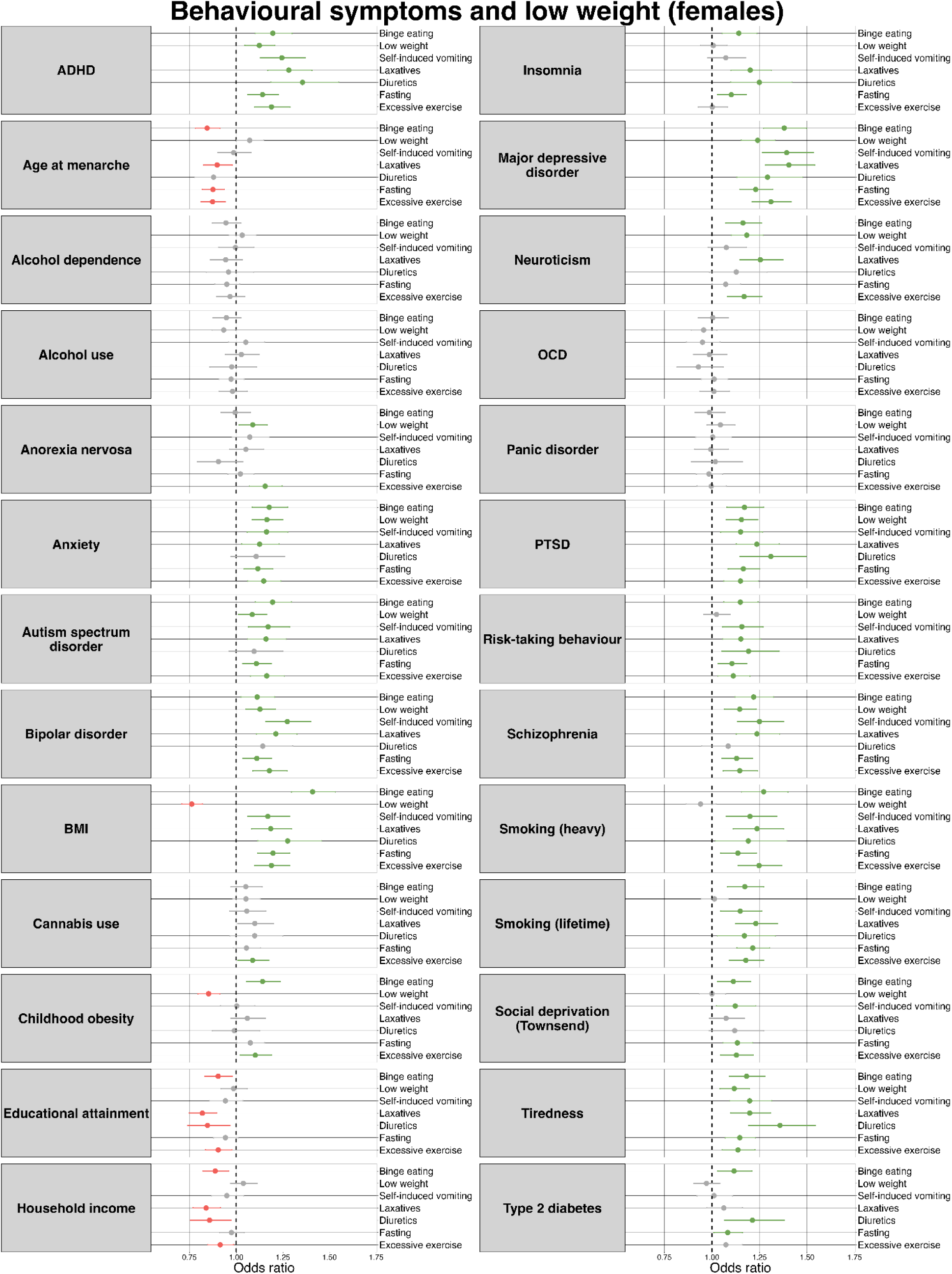
Anthropometric, psychiatric, social, behavioural, somatic, and personality polygenic risk score associations with lifetime behavioural eating disorder symptoms and low weight in females.

Three PGSs were negatively associated with age at onset in females (**Supplementary Figure 5; Supplementary Table 18**) and none in males (**Supplementary Table 19**). In females, higher childhood obesity PGS was associated with a younger onset of binge eating (beta = -0.56, 95% CI -0.82, -0.31, *q* = 0.003); with each SD increase in the PGS corresponding to, on average, a ∼7 months earlier onset. Similarly, each SD higher educational attainment PGS was associated with an onset of low weight ∼6 months earlier (beta = -0.54, 95% CI -0.81, -0.27, *q* = 0.006) whilst a one SD higher schizophrenia PGS was associated with an earlier onset of fasting by ∼7 months (beta = -0.56, 95% CI -0.83, -0.29, *q* = 0.003).

We identified several significant PGS associations with cognitive symptoms and their severity in females (**Supplementary Table 20 and 21; Supplementary Figure 6 and 7**) and none in males (**Supplementary Tables 22 and 23**). In females, higher age at menarche PGS was associated with lower odds of being afraid of weight gain (OR = 0.88, 95% CI 0.81, 0.95, *q* = 0.01) and feeling fat at low weight (OR = 0.89, 95% CI 0.83, 0.96, *q* = 0.02). Further, whilst a greater autism spectrum disorder PGS was associated with lower severity of feeling body parts to be larger than they are (OR = 0.9, 95%CI 0.85, 0.96, *q* = 0.01), a greater insomnia PGS was associated with significantly greater severity of self-worth being contingent on body weight or shape at low weight (OR = 1.1, 95%CI 1.04, 1.17, *q*= 0.02), i.e., for each SD higher insomnia PGS, the odds of being in a group that reports greater severity versus all groups reporting less severity combined was 0.1 odds higher, assuming proportional odds across thresholds.

## Discussion

### Phenotypic analysis: Age at onset

We extended international research by examining ages at onset for specific eating disorder symptoms in a UK sample. Median onset was 17 years for both fasting and excessive exercise, 16-20 years for purging behaviours (vomiting, laxatives, diuretics); and 18 and 20 years for binge eating and low weight. Our UK findings align with international estimates that eating disorders typically emerge between adolescence and early adulthood (7). Despite this existing research, there is a stereotype that eating disorders are primarily adolescent conditions. Our findings challenge this by showing onset is equally common in adulthood.

Our findings suggest that limited funding in adult services may disproportionately disadvantage men with eating disorders. Across all symptoms, a higher proportion of males than females reported adult onset (>18 years): binge eating (71% vs. 55%), low weight (81% vs. 57%), vomiting (57% vs. 34%), laxatives (75% vs. 48%), diuretics (81% vs. 53%), excessive exercise (80% vs. 40%), and fasting (82% vs. 42%). Male eating disorders remain under-recognised, under-researched, and shaped by female-centric diagnostic models (51), making inadequate adult service investment especially disadvantageous for men. These patterns may reflect genuine sex differences in onset, driven by pubertal timing, hormonal changes (52) or differential social and environmental risks. Alternatively, they may result from stereotypes framing eating disorders as female conditions (53), leading men to delay recognising symptoms and report later onset (54). Further research using longitudinal prospective research is needed to clarify these mechanisms.

We demonstrate the need for parity between child/adolescent and adult services in the UK. Historically, adult services have experienced greater constraints on funding and resources, with 55% of child/adolescent services reporting the discharge of patients at age 18 for not meeting the criteria for adult services (55). Stricter thresholds reflect limited resources. In 2014, the Children and Young Person’s community eating disorders services in England were allocated an additional £30 million (now ∼£53 million) per year (56). Adult services saw no equivalent increase until the 2019 NHS Long Term Plan (57), which outlined investment in adult community eating disorder services to improve the continuity of care (58). Beyond different therapeutic approaches, transitions are hindered by poor communication, unclear guidelines, and disruption of established therapeutic relationships at a critical developmental stage (59–61). Our results support not only sustaining but expanding this investment.

### Genetic analysis: SNP-based heritability

We found that whilst each lifetime symptom has a significant genetic component (0.31–0.78), their age at onset appears genetically influenced. Our null findings may reflect limited sample size; Harder et al. (2022) reported a 5.6% heritability for depression onset in 76,365 participants (21), while Ferentinos et al. (2015) found 17% in 3,468 participants using structured interviews. Interview-based phenotyping may yield more reliable data than surveys (62,63). Further, other quantitative phenotypes have demonstrated low heritability estimates, such as subjective well-being (h^2^ = 0.062 [SE = 0.005]), cigarettes per day (0.057 [0.013]), and extraversion (0.049 [0.008]) (25). GCTA-GREML heritability estimates are sensitive to measurement error (64), which may be greater in quantitative than binary phenotypes. For example, recalling whether symptoms began at 16 or at 17 is less reliable than before or after 30. Nonetheless, even binary onset phenotypes showed non-significant heritability. Watson et al. (2022) reported 1% heritability for age at diagnosis in anorexia nervosa, with 9,335 participants and mixed phenotyping methods, including clinical interviews. Thus, our null results may reflect both smaller sample size and measurement error.

We found no molecular genetic evidence for heritability of cognitive symptoms or their severity (e.g., fear of weight gain, self-worth contingent on body shape/weight), despite twin studies reporting moderate estimates (e.g., 23% for feeling fat at low weight) (28). As with age at onset, smaller genetic contributions may exist but remain undetectable in our sample. Larger meta-analyses are needed to clarify whether polygenic influences on these symptoms can be reliably identified.

Whilst eating disorder symptoms are heritable, common genetic variation may contribute little to onset timing or cognitive severity. Instead, environmental factors - such as early trauma (65), cultural pressure around thinness, urbanisation, industrialisation (66), consumerism, and changing gender roles (67) - along with non-common genetic factors, may better explain individual differences in these traits.

### Genetic analysis: Cross-trait polygenic associations

In females, we observed that a higher BMI and childhood obesity PGS were linked to lower odds of low weight, while a higher anorexia nervosa PGS was linked with a higher odds. This supports prior findings that combined low-BMI and AN PGSs strongly shape BMI trajectories (68). We also provide the first evidence that greater childhood obesity genetic load is associated with earlier binge eating onset (∼7 months) in females. Higher BMI PGS was positively associated with all behavioural symptoms in females and most in males, and most cognitive symptoms and their severity in females, consistent with prior associations between anthropometric PGSs and eating disorder symptoms such as body dissatisfaction and fasting (69).

The type 2 diabetes PGS was positively associated with binge eating, diuretics, and fasting but not low weight in females, despite type 2 diabetes having a negative genetic correlation with AN (r_g_ = −0.22) (70). Likewise, the OCD PGS was unrelated to low weight, despite the known genetic overlap between OCD and AN (71) and body dysmorphic concerns (72), suggesting metabolic and psychiatric traits may differentially influence low weight and psychiatric symptoms of AN.

PGSs for ADHD and PTSD in females and PGS for MDD in males and females showed positive associations across all symptoms, supporting shared psychiatric genetic risk (73). We replicate links between earlier menarche and binge eating (40), and report - to our knowledge - novel associations with laxative use, fasting, excessive exercise, and feeling fat and fear of weight gain at low weight. Some mechanisms remain unclear: menarche and disordered eating are negatively genetically correlated (r_g_ = -0.18) (74); pubertal fat gain may heighten body dissatisfaction (75); and estrogen may moderate genetic effects on binge eating during puberty (76).

In females, the educational attainment PGS was linked to lower odds of diuretic/laxative use and earlier onset of low weight, but not low weight itself.. This contrasts with prior positive genetic correlations between years of schooling and both early- (r_g_ = 0.15) and typical-onset AN (r_g_ = 0.24) (39). Emerging evidence suggests the non-cognitive genetic component of education (e.g., curiosity, self-control) may drive this link (77). Our results highlight the value of symptom-level analyses for disentangling genetic pathways to eating disorder risk.

### Limitations

Our findings must be considered in light of multiple limitations. First, age at onset was measured by retrospective self-report, which is unreliable (45,78); in our data, 12% of binge eating onset estimates differed by ≥10 years. Question wording also varied. For example, that low weight showed the oldest onset - contrary to other studies (7) - may reflect its measurement as an *outcome* of other symptoms such as restrictive eating and weight loss behaviours. Compensatory behaviours’ first onset were recorded whilst binge eating was defined as clinically-relevant onset (“…*regular episodes of binge eating*”). Whilst self-report enables efficient large-scale data collection, careful question design is important (see (79)). Second, COPING data were collected throughout the COVID-19 pandemic, when environmental homogeneity may have inflated heritability. Third, our sample is largely white, female, and highly educated, with genetic analyses limited to European genetically-inferred ancestry; such selection can bias genotype-phenotype associations (80). Ongoing recruitment efforts in GLAD and EDGI UK aim to increase diversity, particularly among males and other genetic ancestries.

### Future directions

Our work raises questions for future research. The establishment of twin-based heritability estimates for the age at onset of eating disorder symptoms would contextualise our SNP-based heritability estimates (39). Although incorporating age at onset can boost GWAS power, this relies on onset being heritable (21); larger samples and more precise phenotyping are needed to test this in eating disorders.

## Conclusion

To conclude, we show that eating disorders arise in adulthood as frequently as in childhood or adolescence, challenging the stereotype of primarily adolescent onset and carrying important clinical implications. The observed sex differences in age at onset may reflect true biological variation or reporting bias and require further study. While individual eating disorder symptoms are clearly heritable, the timing of their onset and cognitive features appear only minimally influenced by common genetic variation.

## Acknowledgements

We would like to acknowledge all GLAD, EDGI UK, and COPING participants for their time in taking part in our studies.

## Author Contributions

HLD, MH, GB, CH formulated the research question and designed the study; all authors were involved in data collection and cleaning; HLD and CH carried out the analyses for the study, with assistance from ARTK and RW; HLD wrote the article with help from CH and critical review from all authors; all authors gave final approval of the version to be published and agree to be accountable for all aspects of the work in ensuring that questions related to the accuracy or integrity of any part of the work are appropriately investigated and resolved.

## Transparency Declaration

HLD affirms that the manuscript is an honest, accurate, and transparent account of the study being reported; that no important aspects of the study have been omitted; and that any discrepancies from the study as planned (and, if relevant, registered) have been explained.

## Data Availability

The data in this study are available via application to the relevant study.

## Research Material Availability

See data availability statement.

## Relevance statement

Our findings challenge the stereotype that eating disorders are primarily adolescent conditions: onset occurs as often in adulthood, with males more likely than females to report adult onset across all symptoms. This demonstrates the need to strengthen adult services and improve recognition of male presentations. Genetically, while eating disorder symptoms are heritable, the timing of onset and cognitive symptoms showed minimal heritability, which needs further exploration with larger sample sizes and more thorough phenotyping approaches. Cross-trait polygenic associations revealed distinct pathways, underscoring the value of symptom-level assessment in capturing diverse presentations and guiding more tailored, patient-centred care.

