## Supplementary Figures for "Mapping the genetic landscape of the age at onset and severity of eating disorder symptoms"

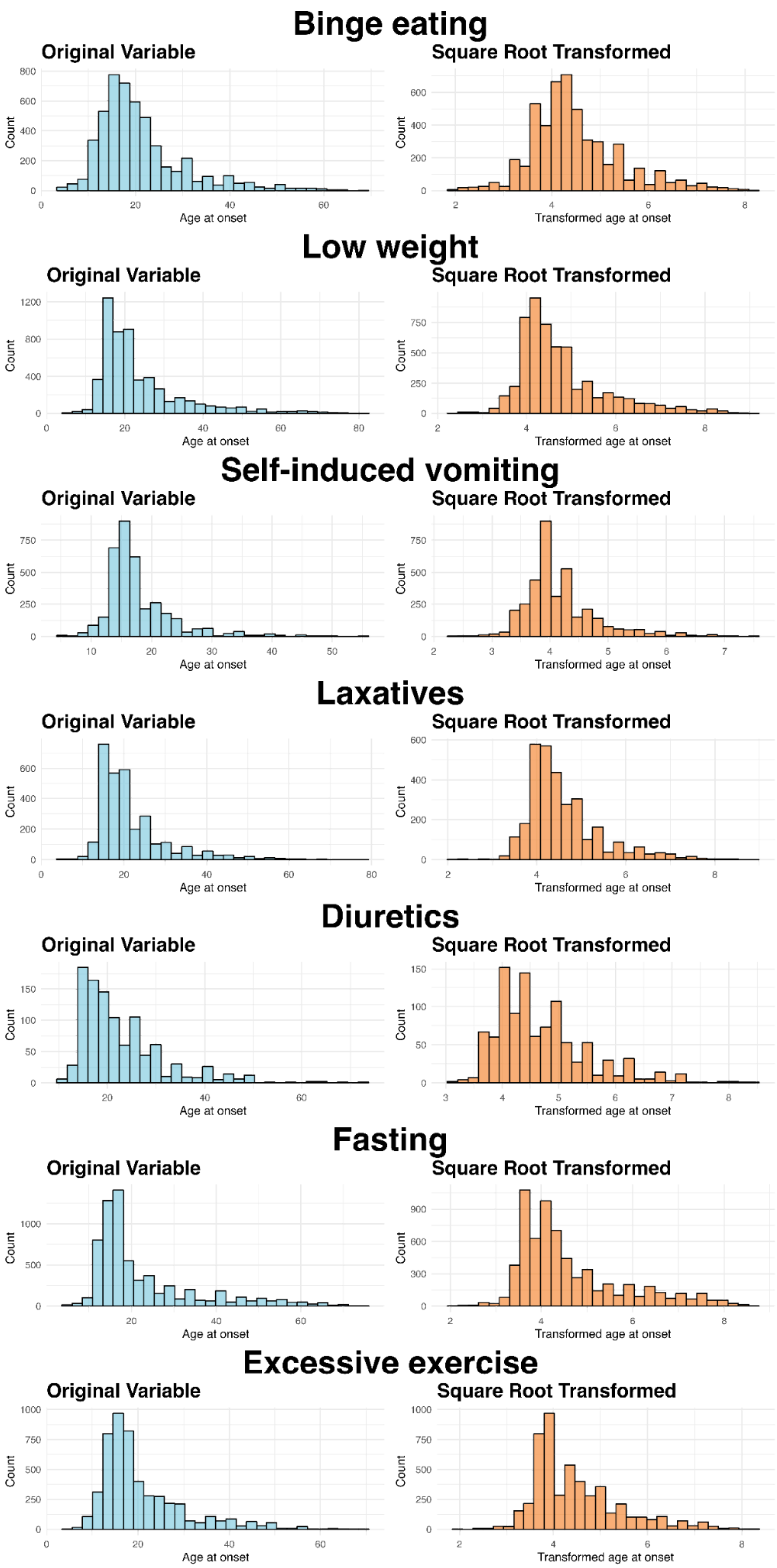


**Supplementary Figure 1.** Original age at onset variable for each behavioural eating disorder symptom and low weight and transformed via square root.

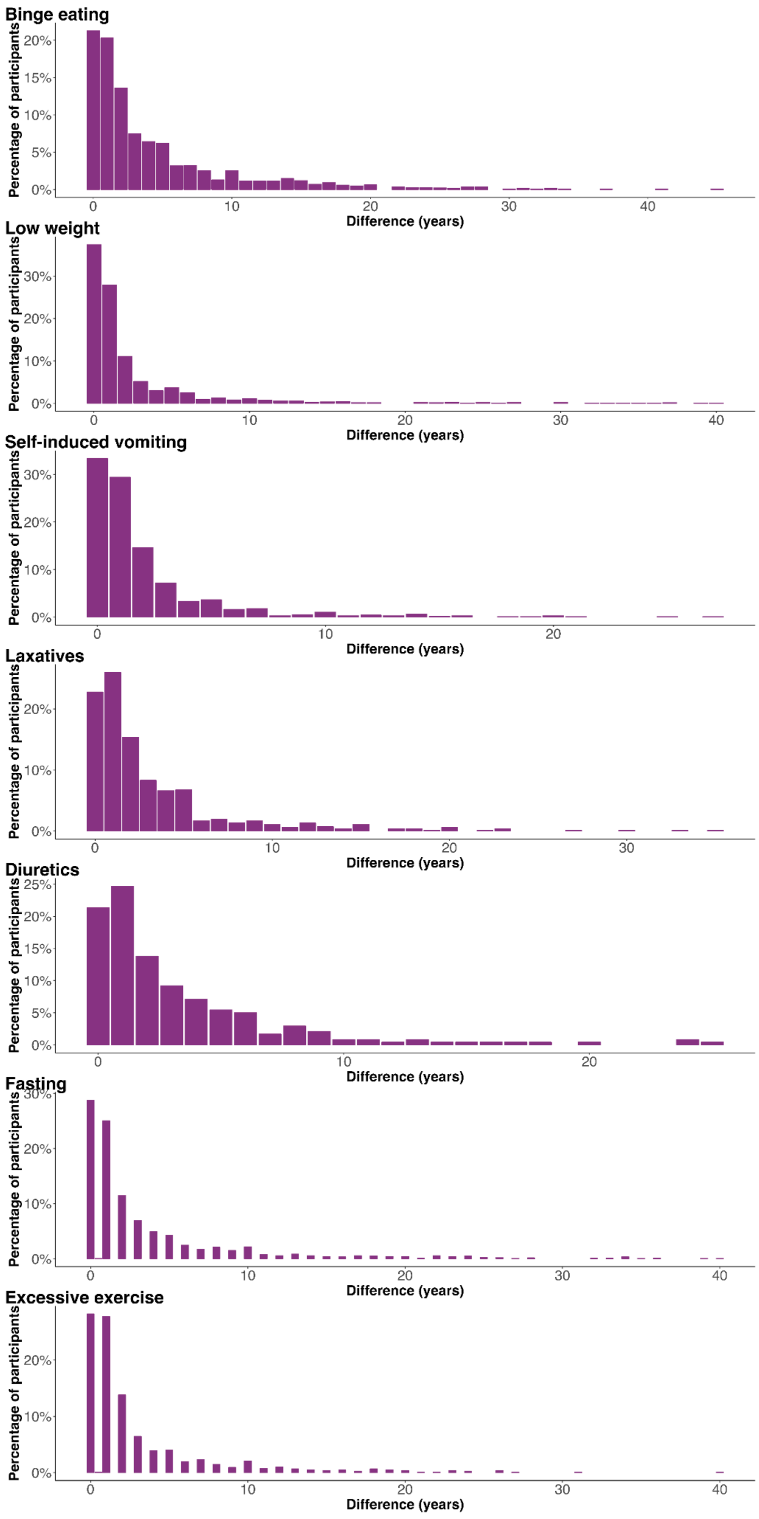


**Supplementary Figure 2.** Differences in reporting of age at onset in the Genetic Links to Anxiety and Depression Study and the COVID-19 Psychiatry and Neurological Genetics study.

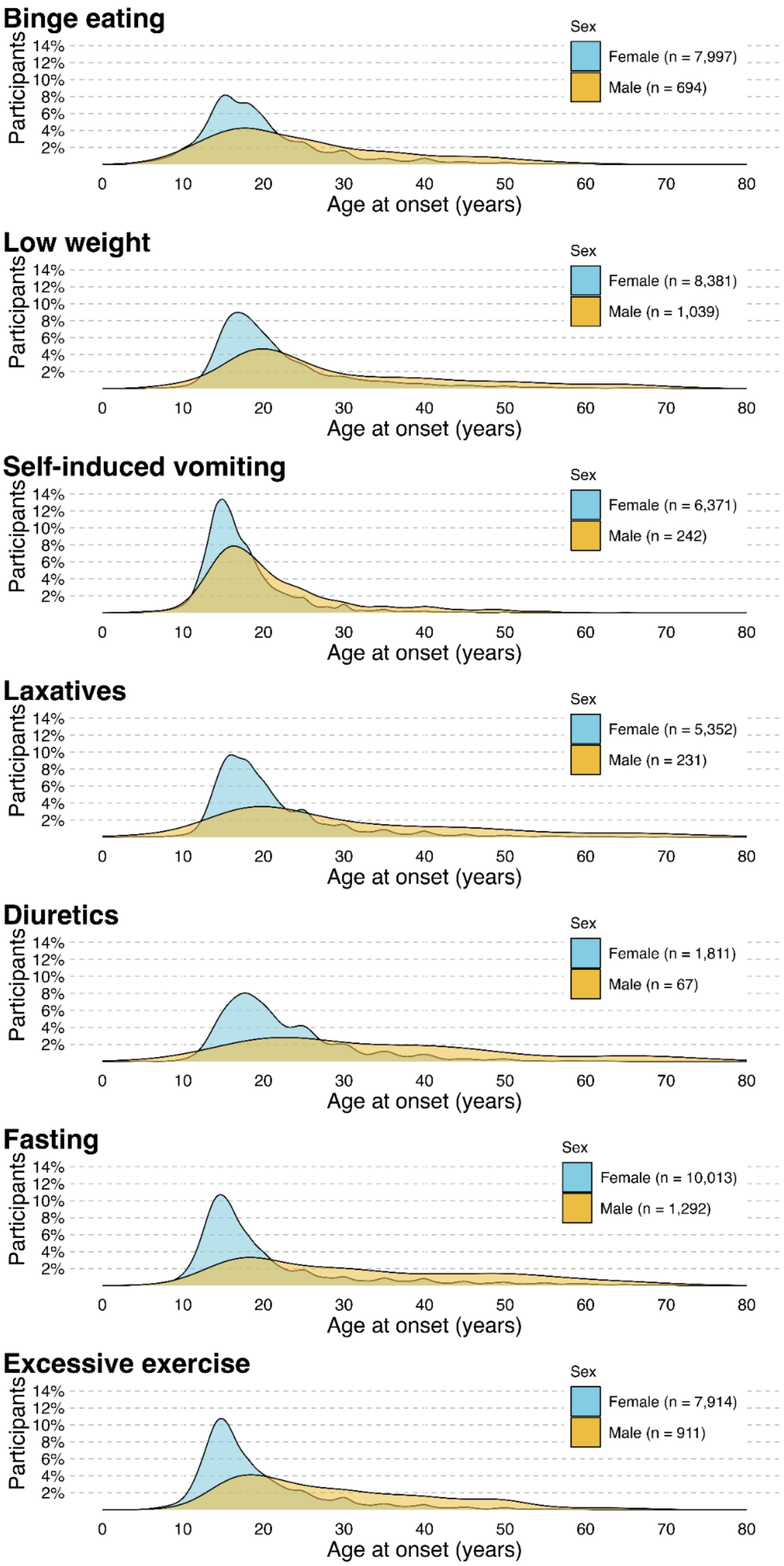


**Supplementary Figure 3.** Sex-stratified age at onset of behavioural eating disorder symptoms and low weight.

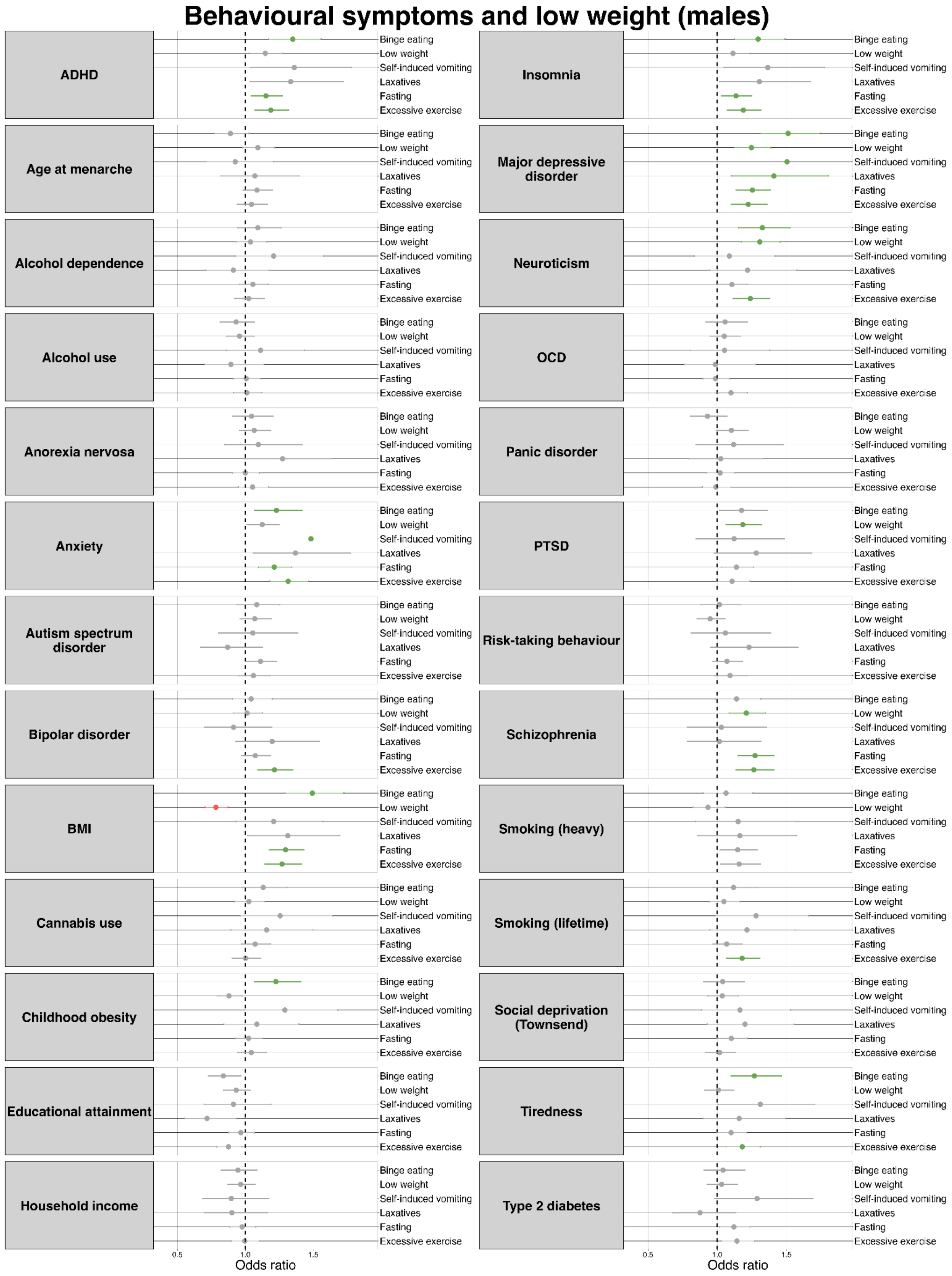


**Supplementary Figure 4.** Anthropometric, psychiatric, social, behavioural, somatic, and personality polygenic risk score associations with lifetime behavioural eating disorder symptoms and low weight in males.

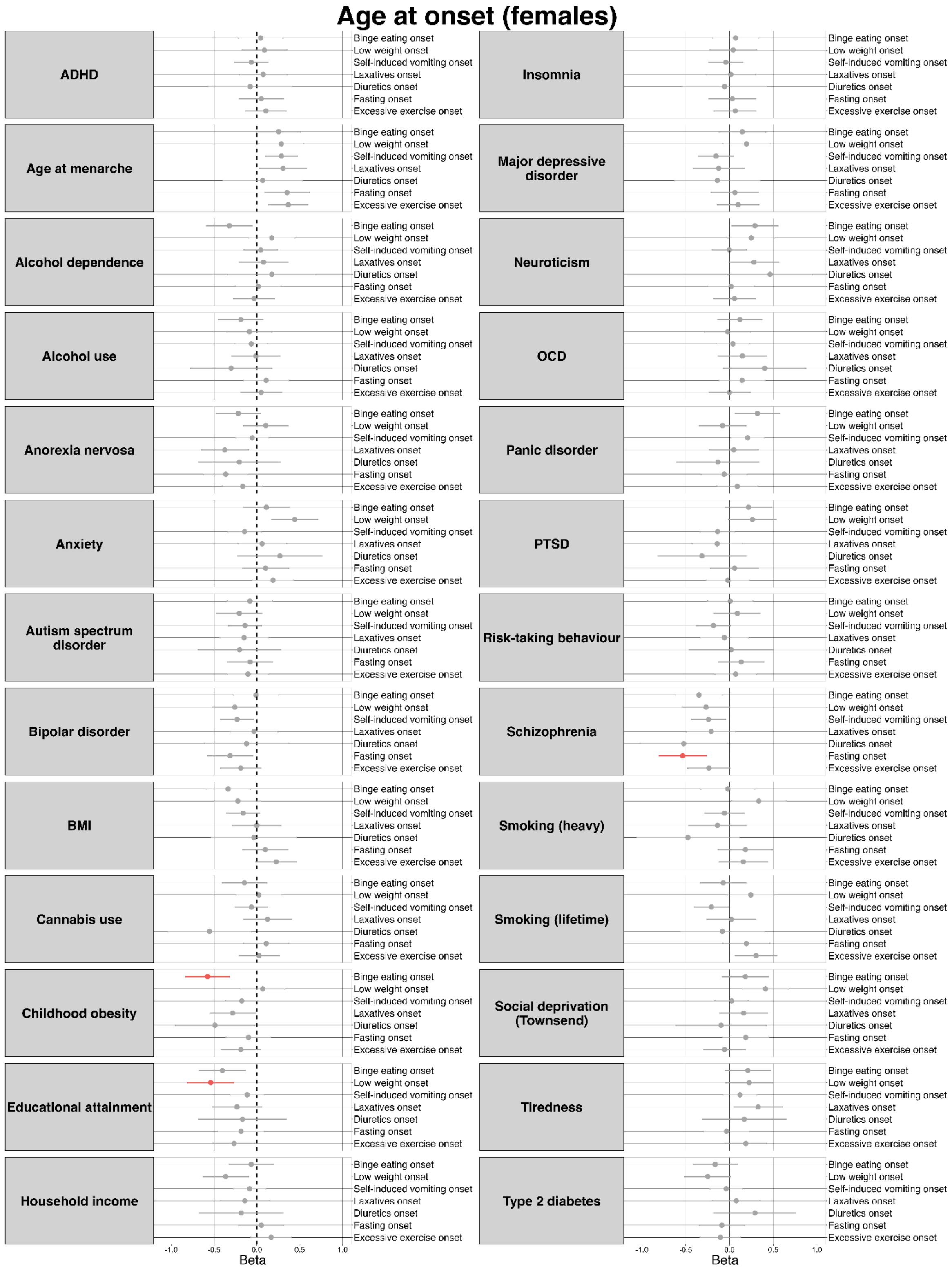


**Supplementary Figure 5.** Anthropometric, psychiatric, social, behavioural, somatic, and personality polygenic risk score associations with age at onset of behavioural eating disorder symptoms and low weight in females.

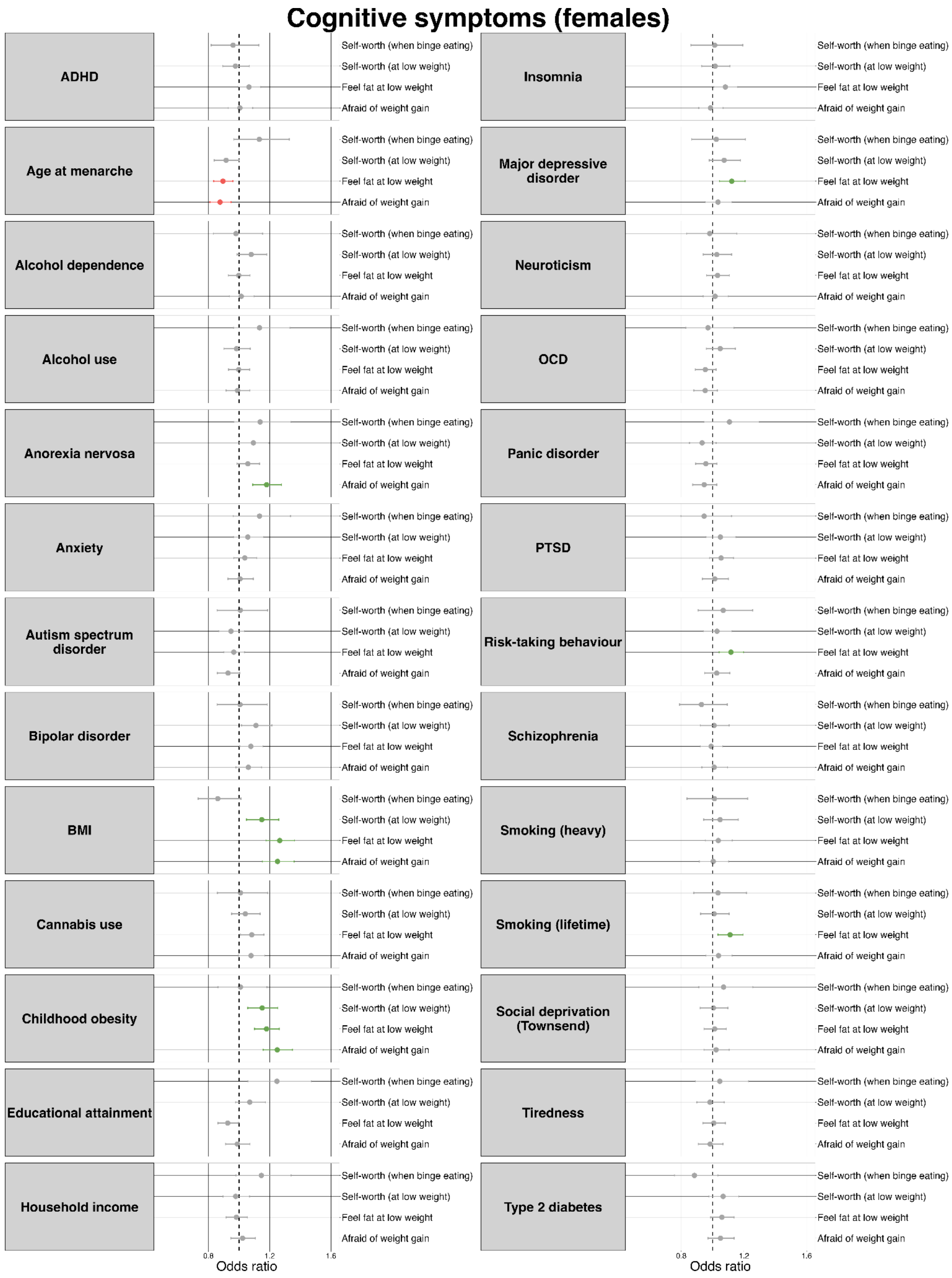


**Supplementary Figure 6.** Anthropometric, psychiatric, social, behavioural, somatic, and personality polygenic risk score associations with lifetime cognitive eating disorder symptoms in females.

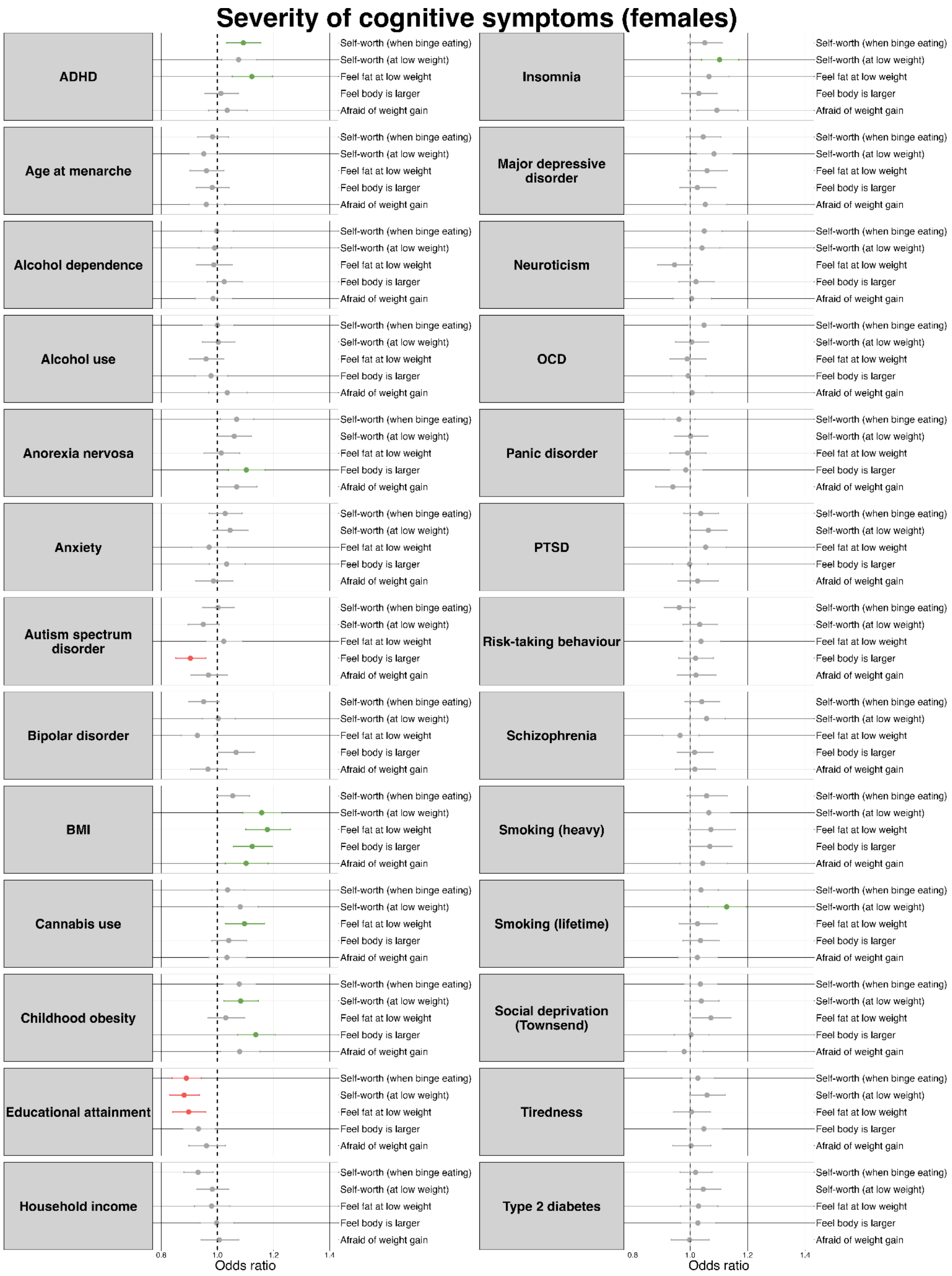


**Supplementary Figure 7.** Anthropometric, psychiatric, social, behavioural, somatic, and personality polygenic risk score associations with severity of lifetime cognitive eating disorder symptoms in females.
